# Trait and state mindfulness modulate EEG microstates

**DOI:** 10.1101/2021.11.22.21266675

**Authors:** D. Zarka, C. Cevallos, P. Ruiz, A. M. Cebolla, M. Petieau, A. Bengoetxea, G. Cheron

## Abstract

The present study aimed to characterize microstate dynamics induced by non-reactive attention underlying mindfulness. Electroencephalogram signals from eighteen trained meditators and a matched non-meditators group were recorded before, during, and after a non-reactive attention meditation or during three resting periods respectively, while they were passively exposed to auditory stimulation. In a multimodal approach, microstate cluster decompositions, personality trait questionnaires, phenomenological ratings, and microstates sources localization were analyzed. Our results revealed that temporal parameters of microstates A and C at rest were negatively correlated to mindfulness traits across all participants. After meditation, the frequency of microstate A and C was decreased while microstate B was of longer duration, in meditators. Source localization analysis revealed that the non-reactive trait effect on microstate C at rest was explained by a modified activity of the salience network (identified by the anterior cingulate cortex, thalamus, and insula), while the non-reactive attentional state effect relied on a contribution of (anterior and posterior) cerebellum during meditation. Our results suggest that decreased microstates A and C reflect decreased mental state reactivity, while the increased microstate B relies on attention stability. These findings strongly encourage more research to assess the use of the microstate temporal parameters as a biomarker of the salience network activity, as well as objectify the brain changes induced by non-reactive attention training.

**HIGHLIGHT:**

- The present study aimed to characterize microstate dynamics induced by non-reactive attention meditation, by the use of multimodal analysis including EEG microstate clusters decompositions, personality trait questionnaires, phenomenological reports, and source localization analysis.
- The occurrence of microstate A, recognized to be related to phonological processing and depressive disorders, was negatively correlated to mindfulness trait and was decreased after non-reactive attention meditation.
- The duration of microstate B, generally associated with the visual system, increases after meditation, in particular in meditators with a high non-reactivity trait.
- Temporal parameters of microstate C, recognized to be related to default mode, were negatively correlated to the non-reactivity trait of meditators and were decreased after non-reactive attention meditation. Source analysis revealed that these trait and state effects reflect modified activities of the salient network.

## INTRODUCTION

Ongoing oscillatory brain activities underlie the dynamic patterns of large-scale functional networks. Such patterns emerge even in the absence of an external afferent volley and shape the brain states [McCormick et al., 2020]. They rely on mechanisms of synchronization that orchestrate the activity of widely distributed neuronal populations which reconfigure dynamically to support ongoing cognition and behavior [Buzsáki, 2004]. Such cerebral dynamic patterns are known to produce scalp potential topographies measurable through electroencephalogram (EEG), which remain briefly stable (around 40-120 ms) before rapidly switching into a new one [Lehmann et al., 1987]. These discrete stable topographies, generally named *microstates*, are identified from data-driven clustering methods applied to the time series of EEG voltage maps [Michel and Koenig, 2018]. Microstate configurations are relatively homogenous across studies with four to seven classically distinct topographical patterns (generally labeled A to G) explaining about 70% of the recorded data. Simultaneous EEG-fMRI studies have shown that generators of resting microstates correspond to the fMRI hubs of resting-state functional networks which have been previously associated with phonological processing, visual imagery, attention reorientation, and interoceptive processing [Britz et al., 2010; Custo et al., 2017; Yuan et al., 2012].

The dynamic changes of microstate indicate millisecond time scale modifications of the large-scale networks’ orchestration over time. Accordingly, microstate temporal dynamics are assumed to provide a window into the high order integration processes at the brain scale level [Michel and Koenig, 2018]. The most broadly described variables accounting for each microstate are the frequency of occurrence (occurrence, s^-1^), the meantime of stability (duration, ms), the time percent of total explained signal it covers (coverage, %), and the percent of total variance it explains (GEV, %). The temporal parameters of microstates were investigated in a wide range of studies related to conscious processes such as states of alertness [Brodbeck et al., 2012; Comsa et al., 2019; Zanesco et al., 2021a], spontaneous phenomenal experiences [Lehmann et al., 2010; Pipinis et al., 2017], self-generated cognition [Bréchet et al., 2019; Milz et al., 2016; Seitzman et al., 2017], personality trait [Schiller et al., 2020; Zanesco et al., 2020], psychiatric disorders [Damborská et al., 2019; Grieder et al., 2016; Rieger et al., 2016], as well as psycho-behavioral practices [Bréchet et al., 2021; Faber et al., 2017; Katayama et al., 2007; Panda et al., 2016; Zanesco et al., 2021b].

In this context, the present study aimed to investigate microstate dynamics underlying mindfulness meditation. Mindfulness meditations refer to a family of attention-based practices imported from Buddhist contemplative traditions [Dahl et al., 2015]. They rely on the ability to purposely maintain the attention instant-by-instant to the lived experience in a non-reactive stance toward the experience content [Kabat-Zinn, 1990]. These practices are thought to reinforce attention stability and meta-awareness abilities [Lutz et al., 2008]. They are generally divided into focused attention meditation (FA) and non-reactive attention meditation (OM, also called open-monitoring meditation) [Dahl et al., 2015]. FA relies on effortful control of attention (i.e selective attention, sustained focus, shifting attention) [Hasenkamp and Barsalou, 2012] and is frequently used to install attentional stability previously to OM. In contrast, OM relies on effortless, no-directed attention while keeping arousal, emphasizing a non-reactive stance towards experience (openness, vigilance, and non-reactivity) [Lutz et al., 2008]. This non-reactivity has been defined as the tendency to allow thoughts and feelings to come and go, without getting caught up in or carried away by them [Baer et al., 2008]. This promotes an inhibitory state on the body and mind movement and attenuates the weight of emotional and cognitive habits to favor the ongoing awareness of the present experience [Fucci et al., 2018; Zorn et al., 2021]. It has been notably shown that OM is associated with a reduced attentional blink [van Vugt and Slagter, 2014], and changes in acoustic startle reflex intensity [Antonova et al., 2015; Levenson et al., 2012]. Further, consistent meditation training has been associated with long-term psychological changes that can be assessed as mindfulness traits through various questionnaires [Baer et al., 2006; Baer et al., 2008; Brown and Ryan, 2003].

Attentional-based meditations are known to engage several large-scale networks including the central executive network (including the attentional networks), the silence network (SN), and the default mode network (DMN) [Lutz et al., 2015]. However, most of the previous meditation studies focused on FA, and little is known about the OM-induced brain dynamics. Regarding microstate studies, in particular, a recent longitudinal study has indicated that 6-week digital training of breath-focused meditation-induced topographical changes in microstate clustering collected at rest in novice meditators [Bréchet et al., 2021]. Source localization methods showed that these topographical changes were supported by the modified activity of the insula, supramarginal gyrus (BA40), and superior frontal gyrus (BA10) [Bréchet et al., 2021]. Moreover, increased occurrence and duration of DMN-related microstates (generally referred to as C) have been reported at rest in expert meditators compared to the control group, and also in expert meditators during FA meditation compared to rest. Interestingly, these dynamic changes were associated with decreased PCC connectivity in meditators compared to controls, which further decreased during FA meditation [Panda et al., 2016]. In contrast, transcendental meditation, an FA-like meditation closer to OM practices [Travis and Parim, 2017], brings internally generated thoughts detachment which has been related to the lower occurrence of microstates A and C, linked to phonological and interoceptive processing respectively [Faber et al., 2017]. 3 months of full-time meditation training involving both FA and OM, resulted in attentiveness and serenity increases, as reported by participants, which have been associated with microstates duration reduction at rest [Zanesco et al., 2021b].

In the present study, we focused on microstates dynamics induced by OM in participants after a standardized 8-week training. We hypothesized that the non-reactive state underlying OM practices induce changes in temporal dynamics of microstates, and more particularly they should reduce the temporal feature of the microstate C which has been linked to the salience network and the default mode network [Britz et al., 2010; Michel and Koenig, 2018]. Concretely, we compared EEG signals recorded before, after, and during OM in the trained participants, with those recorded at rest during equally, three periods of time in a matched control group of waitlist participants. During recordings, regular auditory stimuli were passively displayed in the background environment to assess participants’ reactivity to distractors. We investigated microstate dynamics induced by OM dissociating the related state from trait effects.

In line with previous studies, we expected to reveal the four to seven classical microstates. We hypothesized that (1) trained meditators should show different temporal characteristics of microstates compared to non-meditators [Zanesco et al., 2021b]. In particular, OM should induce microstate C temporal parameters to decrease compared to the rest [Faber et al., 2017]. We also hypothesized that (2) microstate dynamics will be related to mindfulness traits and phenomenological aspects of experience. Additionally, (3) we questioned change previously reported in microstate topographies between groups induced by meditation training [Bréchet et al., 2021].

## MATERIALS AND METHODS

### PARTICIPANTS

Forty healthy volunteers (aged 25 to 65 years) were recruited during informative sessions of standard MBSR programs provided by the continuing education center of the *Université Libre de Bruxelles* (ULB HELSci, Brussels, Belgium). Exclusion criteria were having significant experience of mindfulness meditation, history of epilepsy, hearing troubles, attention deficit with/without hyperactivity, history of substance abuse, and antecedent of psychiatric disorder. All participants provided written consent after a full explanation of the investigation. This study was approved by the ethics committee of the academic hospital Erasme (Brussels, Belgium), in agreement with the Belgian law relative to research on humans [Dresse, 2005] and the Helsinki declaration [World Medical Association, 2013].

20 among the participants, followed an 8-week meditation training (MED, 10F/10M, mean age: 41.68 ±10.91 years) and the 20 others constituted a waitlist matched control group (noMED, 11F/9M, mean age: 43.50 ±15.76 years). The meditation training was dispensed by senior instructors certified by the Center for Mindfulness (UMass Medical School, Massachusetts, USA) with more than 15 years of mindfulness teaching expertise. They followed the standard structure of the mindfulness-based stress reduction program [Crane et al., 2017; Kabat-Zinn, 1990] respecting the teaching assessment criteria [Crane et al., 2013]. Following these standards, the training program was composed of a 3-hour intensive group session per week for 8 weeks, plus one full day (7 hours, at week 6) intensive group session. The first 4 weeks emphasized attention stability reinforcement (FA), while the following 4 weeks (including the full day of practice) emphasized meta-awareness (OM). Participants were further encouraged to practice at home for 45 min daily during the 8 weeks using provided audio files. To assess participants’ commitment, they were instructed to report each day the type of exercises and the time spend to practice using a standardized table. One subject was unable to finish the training for personal reasons and was excluded from the study. Accordingly, the meantime of daily practice per participant was 23.41 (±11.25) min, totalizing a mean of 52.84 hours (±11.35h) of practice per participant during the training.

### PERSONNALITY TRAIT ASSESSMENT

Participants engaged in the training program were asked to complete the French version of the Five Facets Mindfulness Questionnaire (FFMQ) [Baer et al., 2006; Heeren et al., 2011] before and after training. The FFMQ is a commonly used assessment of mindfulness traits showing good reliability and validity [Baer et al., 2008; de Bruin et al., 2012; Carmody and Baer, 2008; Heeren et al., 2011; Soler et al., 2012]. Its specificity is to be constructed as a multidimensional assessment based on five previous scales allowing to characterize mindfulness according to five sub facets: observing, non-reactivity to inner experience, non-judgment, describing and acting with awareness [Baer et al., 2006; Baer et al., 2008]. FFMQ is responsive to various forms of mindfulness training [Khoury et al., 2013] as well as to the amount and quality of mindfulness practice [Goldberg et al., 2014]. Comparison of FFMQ scores before and after training confirmed a significant increase after the 8-weeks training (paired t-test, FFMQ: t = 4.372, p < 0.001; observing: t = 2.929, p = 0.010; describing: t = 3.528, p = 0.003; non-reactivity: t = 0.331, p = 0.745; non-judgement: t = 4.737, p < 0.001; Acting with awareness: t = 3.378, p 0.004). To allow traits comparison between groups, wait-list participants were also asked to complete the FFMQ a few days before experiments. All participants were asked to complete the Hospital Anxiety and Depression Scale (HAD) [Zigmond and Snaith, 1983], as well as the Perceived Stress Scale (PSS) [Cohen et al., 1983; Nielsen et al., 2016] and the Pittsburgh Sleep Quality Index (PSQI) [Buysse et al., 1989].

### PROTOCOL

#### Recording periods (RS/OM)

Experiments took place in the Laboratory of Neurophysiology and Movement Biomechanics (ULB) in Brussels, Belgium, a few days after the training program. The EEG recording was composed of three eyes-closed periods of 8 minutes each. The first period corresponded to resting-state recordings for both groups (RS1). Participants were instructed to keep their eyes closed, motionless, to stay awake, and to wait for the end of the recording which was signaled by a bell. During equipment installation brief semi-structured conversation oriented to participants’ daily life was engaged by the experimenter (Supplementary Files 1). The second period corresponded to another resting-state recording for controls (RS2) and a not guided OM for meditators. Following the training program, OM was preceded by FA to stabilize attention and it was introduced by the same standardized oral instruction inviting to open the attentional scope to the experiential field. The non-meditators were instructed as in the first block and they respected the same amount of time as the meditators. The last period (RS3) corresponded to a new period of resting-state for both groups, with similar instructions as the first block.

#### Phenomenological assessment

Concomitantly, auditory stimuli (540/440Hz, 80 dB, 101 ms duration, ISI: 3s) were presented via loudspeakers bilaterally placed behind the subject during the whole EEG recording. To avoid the surprise effect, participants were exposed to the stimuli for 30 seconds before recordings. They were instructed to not pay attention to them. At the end of each recording, participants were asked to rate on a 5-point Likert scale (1 = not present at all; 5 = extremely present) about their auditory distractibility, mind wandering, emotional charge, body discomfort, and sleepiness during recording. Questions were formulated as *“Please rate how tired you were”* according to Brandemeyer et al. [2018] (Supplementary Files 2). Short structured interviews (adapted from Petitmengin et al. [2019]) oriented on the description of their experience according to each phenomenal aspect were also made. Reports were compared with a standardized criteria-based scale to refine rating across participants (Supplementary Files 2). The experimenter performing the interviews was blinded regarding groups.

#### EEG SETUP AND PREPROCESSING

EEG recordings were made using the ASA system (ANT software, the Netherlands) with 128 Ag/AgCI sintered ring electrodes embedded in an active-shield cap (10-20 system) and shielded co-axial cables. Eye movements were recorded using two bipolar electrodes: one placed horizontally on each outer eye canthus, the other placed vertically above and below the right eye. All electrodes were referred to the ears lobes. The ground electrode was placed in the neck on the C7 spinous process. Impedances were kept below 10 kΩ and checked before each block recording. Signals were recorded with a sampling rate of 2048 Hz and a resolution of 16 bits. We used EEGLAB software [Delorme and Makeig, 2004] for offline data treatment. Data were band-pass filtered between 1 and 40 Hz. Filtered EEG was down sampled to 125Hz. Noisy electrodes (max 10%) were interpolated using three-dimensional spherical splines. Cleaned EEG was re-referenced to the average reference. EEG data was reduced to 110 channels to remove muscular artifacts originating in the neck and face. Six minutes of continuous EEG data per block were selected for the analysis. At this stage, data from three subjects (1 MED, 2 noMED) were rejected due to large artifacts. The final analysis concerns 36 subjects (18 MED, 10F/9M, mean age: 42.39 ±10.78 years; 18 noMED, 10F/8M, mean age: 43.22 ±16.02 years)

### MICROSTATES ANALYSIS

Microstate analysis was performed with freely available Cartool Software 3.70 [Brunet et al., 2011]. Following standard procedures [Michel and Koenig, 2018], we used k-means clustering to estimate the set of topographies explaining the EEG signals for each participant and each period. Only data points at local maximum GFP (global field power) were considered for clustering to improve the signal-to-noise ratio. The polarity of the maps was ignored. The cluster maps of each participant were subjected to a second k-means cluster analysis across participants to estimate the set of topographies explaining the EEG signals for each group and period. To distinguish trait differences between groups and state differences between periods, we performed two separate, trait and state, streams of cluster analysis. We first applied the *k*-means clustering on the first resting-state recording (RS1) data across all participants (accordingly to Damborská et al. [2019] methods) to investigate the trait effect between groups. We used six independent optimization criteria merged in a single metacriterion to determine the optimal number of clusters [Pascual-Marqui et al., 1995; Bréchet et al., 2019]. We then applied the *k*-means clustering across participants according to groups and periods (RS1, RS2/OM, RS3) (accordingly to Bréchet et al. [2021]) to investigate the state effect. Here, we fixed the number of clusters for each group and period accordingly to the optimal number of clusters determined by previous trait analysis (Figure 1).

**Figure 1:**
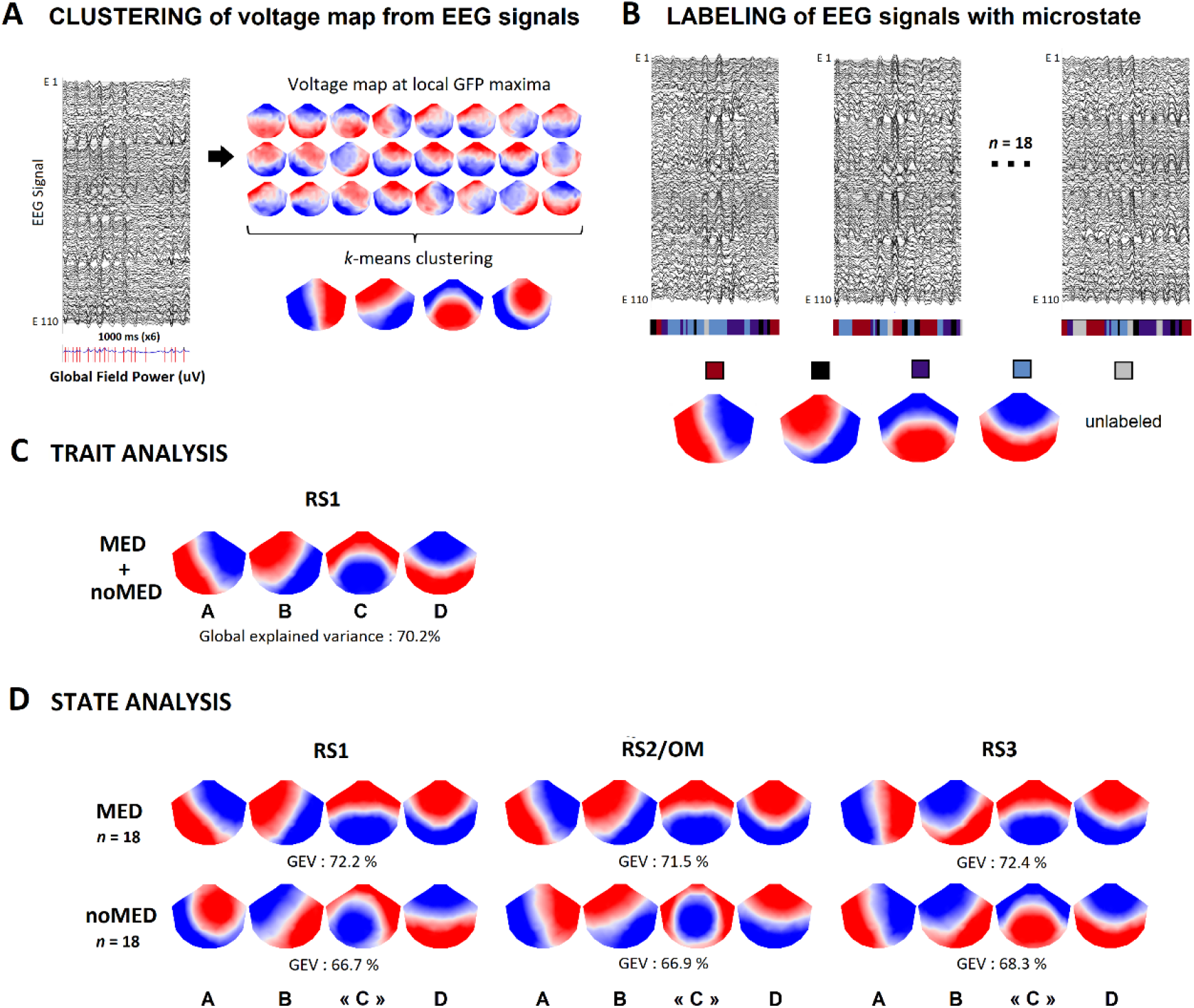
Microstate analysis method. (A) Eyes-closed EEG signals from 110 electrodes. Voltage maps at local GFP peaks are identified as periods of topographic quasi-stability. A first *k*-means clustering determined the subjects-level clusters from voltage maps at local GFP peaks; (B) After a second *k*-means clustering identifying global clusters from subjects-level microstates, the original EEG signals from each recording are continuously labeled with the best-correlated global microstate map. The occurrence, duration, and coverage for all microstates were calculated from these labeled EEG recordings; (C) Set of four cluster maps best explaining the data as revealed by *k*-means clustering across all participants (meditators and non-meditators) at first 8-min eyes closed recording (RS1); (D) Set of four cluster maps identified by predetermined *k*-means clustering explaining the data according to groups (MED, noMED) and successive periods (RS1, RS2/OM, RS3). Note that microstate labeled « C » showed different topographies between periods within non-meditators (mean spatial correlation: 79.3 ±4.5%) and between groups in RS1 (69.1%) and RS2 (45.1%). These distinctive topographies precluded statistical comparisons of microstate temporal parameters between periods within the non-meditator group, and between groups.

For both streams of cluster analysis, each microstate map identified at the group level was spatially back-correlated (ignoring polarity) with the GFP-normalized map of each participant individually at each data point of their original recording. Thus, each time point of the participant’s original EEG was labeled with the microstate map with the highest correlation. EEG samples that had low spatial correlations (< 0.5) with all group-level microstates were left unassigned. To avoid artificially interrupting temporal segments of stable topography by noise during low GFP, we used a sliding window (half size = 3; Besag factor = 10) [Brunet et al., 2011]. All time points with the same label were then averaged to calculate the four mean maps per participant representing the four microstates. For each clustering decomposition, we calculated the GEV (global explained variance) as the sum of the microstates explained variance (weighted by the GFP at each moment in time) to assess the representativeness of the microstate composition in regards to the original EEG data (Figure 1). We then calculated the duration, occurrence, and coverage of each microstate for each participant and period. The duration is the average time (ms) of contiguous samples labeled according to a specific microstate. The occurrence is the average times per second (s^-1^) a given microstate occurs in the continuous-time series. The coverage is the percent of the total time of the whole EEG recording (%), which was labeled according to a specific microstate [Michel and Koenig, 2018].

### SOURCE ANALYSIS

To estimate the neuronal source accounting for topographical dissimilarities of microstate between groups and periods, we used standardized weighted low-resolution electromagnetic tomography analysis (swLORETA, ASA Software, ANT Neuro, the Netherlands) [Cebolla et al., 2011; Cebolla et al., 2016; Cebolla et al., 2017; Leroy et al., 2017; Palmero-Soler et al., 2007; Zarka et al., 2020; Zarka et al., 2021]. Derived from the sLORETA method [Pascual-Marqui, 2002; Pascual-Marqui et al., 2002; Wagner et al., 2004], swLORETA models spatially distinct sources of neuronal activity from EEG signals without prior knowledge about the anatomical location of the generators even when two dipoles are simultaneously active, and permits the reconstruction of surface and deep sources incorporating a singular value decomposition-based lead field weighting the varying sensitivity of the sensors to current sources at different depths [Cebolla et al., 2011; Cebolla et al., 2016; Palmero-Soler et al., 2007].

We characterized the generators of each mean map per participant and period corresponding to the best spatial correlation with the given microstates on periods of 50 ms. Following standards, the current density of every voxel was divided by the mean current density value of all voxels for every participant and period. This gave us a normalized inverse solution in which a voxel value greater than 1 indicates greater activity than the mean. We then calculated the average of such normalized inverse solution in each period for both groups. To compare groups and periods, we created an image resulting from subtracting the modulus of the swLORETA solution of one group or period from the modulus of the swLORETA solution of compared group or period.

The swLORETA solution was obtained using a 3D grid of 2030 points (or voxels) that represented possible sources of the signal. Based on the probabilistic brain tissue maps provided by the Montreal Neurological Institute [Collins et al., 1994], the solution was restricted to the gray matter and cerebellum. The 2030 grid points (10.00 mm grid spacing) and recording array (128 electrodes) were indexed by the Collins 27 MRI produced by the MNI [Evans et al., 1993]. The Boundary Element Model was used to solve the forward problem [Geselowitz, 1967]. The final coordinates (x,y,z, Talairach coordinates) were obtained using ASA software and identified as Brodmann areas based on the Talairach atlas [Lancaster et al., 2000].

### STATISTICAL ANALYSIS

For trait analysis, we investigate associations between microstate dynamics and personality traits through Spearman’s rank correlation coefficients between temporal parameters (duration, occurrence, and coverage) of each microstate identified by the k-means clustering across all participants for RS1 and scores related to questionnaires (FFMQ, HADS, PSS, PSQI) as well as scores related to phenomenological ratings (mind wandering, auditory distractibility, emotion charge, bodily discomfort, sleepiness). The microstate temporal parameters and the questionnaire scores were also submitted to independent t-tests with Group (MED, noMED) as a between-subject variable. To correct statistics for multiple comparisons, we applied the false discovery rate method [Benjamini and Hochberg, 1995; Lindquist and Mejia, 2015] provided by Cartool Software [Brunet et al., 2011].

For state analysis, scores related to phenomenological ratings were analyzed through Friedman and Mann–Whitney *U* tests with Period (RS1, RS2/OM, RS3) and Group (MED, noMED) as within and between-subject variables. Microstate temporal parameters were submitted to a repeated measure ANOVA according to Period (RS1, RS2/OM, RS3) and Group (MED, noMED). If needed, sphericity violations were corrected by Greenhouse-Geisser correction. Holms corrected paired *t*-tests were used as posthoc tests. We also calculated within the groups, Spearman’s correlation between changes in microstate temporal parameters, phenomenological ratings, and FFMQ scores (and subscales).

For source analysis, the statistical differences between periods were determined by non-parametric corrected permutations as proposed by Nichols et al. [2002] which uses the data itself to generate the probability distribution for testing against the null hypothesis and controls for the false positives that may result from performing multiple hypothesis tests [Nichols and Holmes, 2002]. To perform the permutation, we used the *t*-test as the value of merit. We compute the T-image (T value per voxel) by performing a one-sample t-test (one-tailed) for each voxel of the source space. The null hypothesis is that the distribution of the voxel values of the different images had a bigger mean in one group/period than the other (and inversely). Instead of assuming a normal distribution to assess the statistical significance of the T-score at each voxel, we used the permutation method to create an empirical distribution as explained in detail by Cebolla et al. [2011]. The Holmes maximal correction was calculated separately for each comparison between groups and periods. The 95th percentile of the permutation distribution was used for the corrected maximal statistics which defines the 0.05 level of the significance threshold. Thus, we rejected the null hypothesis for voxel of the unpermuted T-image with *t*-values greater than the 95 ^th^ percentile of the permutation distribution of the corrected maximal statistics [Holmes et al., 1996].

## RESULTS

### QUESTIONNAIRES SCORES

Although no group difference was highlighted for FFMQ total score (FFMQ: *t*(34) = 0.996, *p* = 0.326), we observed that meditators showed significant higher scores for observing (MED: 31.47 ±2.60, noMED: 27.06 ±6.13; *t*(34) = 2.744, *p* = 0.010) and non-reactivity to inner experience facets (MED: 24.53 ±3.18, noMED: 20.28 ±6.20; *t*(34) = 2.528, *p* = 0.016) compared to non-meditators. Other facets (describing, non-judgment and acting with awareness) did not showed significant difference between groups (all *t* < 2.00, *p* > 0.05). Also, our groups of meditators and non-meditators did not differ for sleep quality (PSQI: *t*(34) = -0.054, *p* = 0.958), perceived stress (PSS: *t*(34) = -1.007, *p* = 0.321) as well as anxiety and depression scores (HAD: *t*(34) = -0.348, *p* = 0.348; Anxiety: *t*(34)= -0.639, *p* = 0.527; Depression: *t*(34) = -0.907, *p* = 0.371) (Supplementary Files 3, Table A for details).

### PHENOMENOLOGICAL RATINGS

Ratings showed no period effect on mind wandering and auditory distractibility in non-meditators (both *X*^2^ < 0.130, *p* > 0.937). In contrast, meditators reported lower mind wandering during OM compared to the two other periods (*X*^2^ = 13.298, *p* < 0.001; RS1: *z* = -2.808, *p* = 0.005; RS3: *z* = -2.767, *p* = 0.006), suggesting a successful commitment in meditation practice. They also reported lower auditory distractibility during RS3 compared to RS1 (*X*^2^ = 8.600, *p* = 0.014; *z* = -3.000, *p* = 0.003). Group comparisons showed that meditators reported lower mind wandering during RS2/OM (*z* = -3.616, *p* < 0.001), as well as lower auditory distractibility during RS2/OM and RS3 (RS2/OM: *z* = -1.991 *p* = 0.046; RS3: *z* = -2.785 *p* = 0.010) compared to non-meditators. In addition, and regarding other phenomenal aspects controlled (Supplementary Data 4, 5 for details), group comparisons showed that meditators reported lower emotional charge in RS1 and RS2/OM (*z* = -3.080, *p* = 0.004; *z* = -2.269, *p* = 0.023, respectively) than non-meditators.

### TRAIT MICROSTATE ANALYSIS

Figure 1C illustrates the set of the most dominant topographies obtained by the meta-criterion of the clustering decomposition for RS1 across all participants. The k-means clustering determined that the optimal number of the microstate composition was four (A, B, C, D) explaining 70.18% of the global variance. The four microstate maps corresponded to the classical microstates labeled as A, B, C, and D in literature (Michel, 2018).

Microstate A occurrence and coverage were negatively associated with FFMQ total score (ρ(34) = -0.363, p = 0.032 and ρ(34) = -0.335, p = 0.049, respectively). These correlations were more particularly sustained by the non-judgment (ρ(34) = -0.374, p = 0.027 and ρ(34) = -0.420, p = 0.012 respectively) score of FFMQ. Trends were also observed for non-reactivity score (ρ(34) = -0.333, p = 0.051 and ρ(34) = -0.325, p = 0.057 respectively).

Microstate C duration and coverage were negatively correlated with non-reactivity score of FFMQ (ρ(34) = -0.357, p = 0.035; ρ(34) = -0.352, p = 0.038 respectively). A trend was also observed for occurrence (ρ(34) = -0.325, p = 0.057). Besides, the duration of microstate C was negatively correlated with FFMQ observing score (ρ(34) = -0.388, p = 0.021) and positively correlated with auditory distractibility (ρ(34) = 0.465, p = 0.004) across participants. Auditory distractibility and FFMQ observing score were further negatively correlated (ρ(34) = -0.375, p = 0.026) (Figure 2).

**Figure 2:**
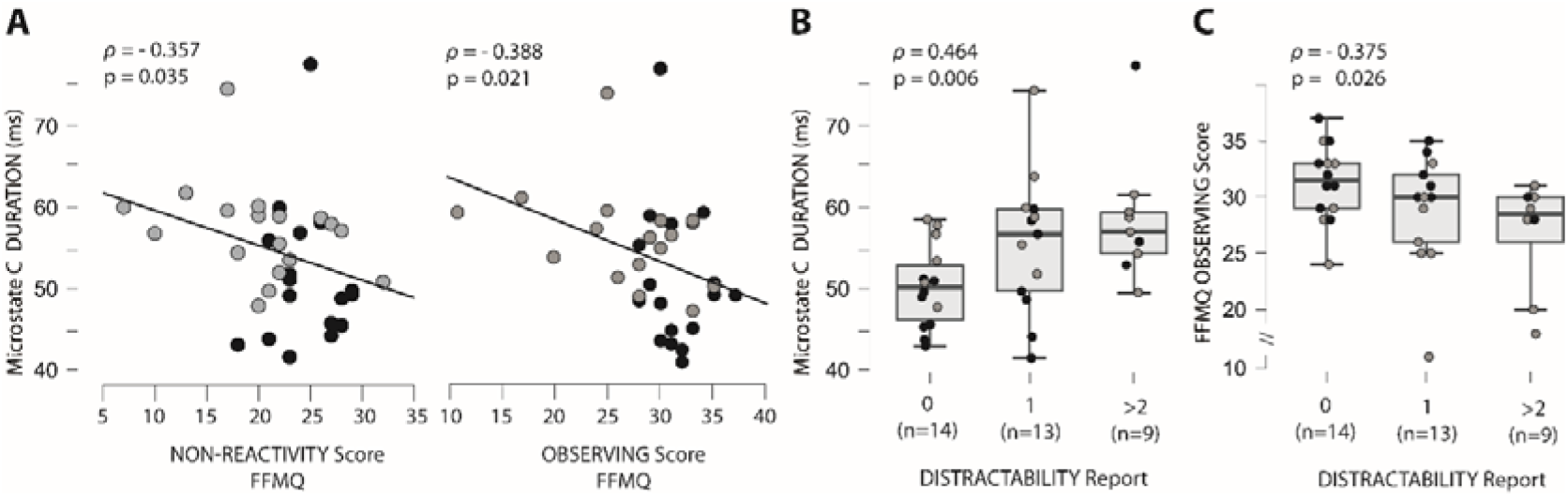
Correlations between microstate C temporal parameters, mindfulness trait, and phenomenological reports across all participants during the first 8-min recording (meditators in black dots and non-meditators in grey dots). We found that (A) duration of microstate C was negatively correlated to score in non-reactivity and observing of FFMQ. Note that meditators showed lower duration of microstate C (*t*(34) = 2.34, *p* = 0.025) as well as higher score in non-reactivity (*t*(34) = 2.528, *p* = 0.016) and observing (*t*(34) = 2.744, *p* = 0.010) than non-meditators. (B) The duration of microstate C was positively correlated to auditory distractibility, while (C) FFMQ observing scores were negatively associated with auditory distractibility ratings.

Post-hoc comparison between group revealed that meditators showed lower occurrence, duration, and coverage of microstate C compared to non-meditators (*t*(34) = 2.16, *p* = 0.038; *t*(34) = 2.34, *p* = 0.025 ; *t*(34) = 2.17, *p* = 0.037 respectively). No significant difference relative to temporal parameters of microstate A, B and D were found between groups (all *t* < 2.00, *p* > 0.05) (Supplementary Files 3).

### STATE MICROSTATE ANALYSIS

Figure 1D illustrates the microstates compositions for both groups and periods RS1, RS2, RS3 for non-meditators and RS1, OM, RS3 for meditators. The maps in meditators were highly similar across periods (RS1, OM, RS2), and they corresponded to the canonical microstates labeled A, B, C, and D in literature (Fig. 1D, upper part). The spatial correlations of these 4 microstates between periods were very high (mean per map: 96.2 ±2.4%). In contrast, non-meditators showed three microstate maps that corresponded to canonical microstates A, B, and D, and another one was less consistent with canonical microstate C, and this, particularly for RS1 and RS2 periods (Figure 1D, lower part). The spatial correlation of these microstates between periods showed a strong correlation for B (96.5 ±1.5%) and D (97.9 ±1.3%), and weaker correlations for A (85,7 ±12.3%) and C (79.3 ±4.5%). Spatial correlation between groups showed strong correlations in the three periods for maps A (94.8 ±6.6%), B (97.7 ±1.4%), and D (96.6 ±2.7%), and a weaker correlation for the fourth map C (69.1 ±24.0%) in particular for RS1 (69.1%) and RS2 (45.1%) periods (RS3: 93.1%). In accord with the weaker spatial correlation concerning the fourth C map, we labeled it « C » across periods in both groups (Figure 1D). The GEV values of each microstate’s composition were 72.2%, 71.5%, 72.4% for meditators in RS1, OM, RS3 periods respectively and 66.7%, 66.9%, 68.3% for non-meditators in RS1, RS2 and RS3 periods respectively. Note that microstate temporal parameters can be only compared between conditions if their microstate compositions present the same topographies [Bréchet et al., 2021; Grieder et al., 2016]. The distinctive topography of microstate « C » precluded comparisons between periods within the non-meditator group, nor between groups. Although we have illustrated the values of non-meditators in graphs for informative purposes, repeated measure ANOVA concerned microstate temporal parameters comparisons between periods (RS1, OM, RS2) within the meditator group only (Figure 3).

**Figure 3:**
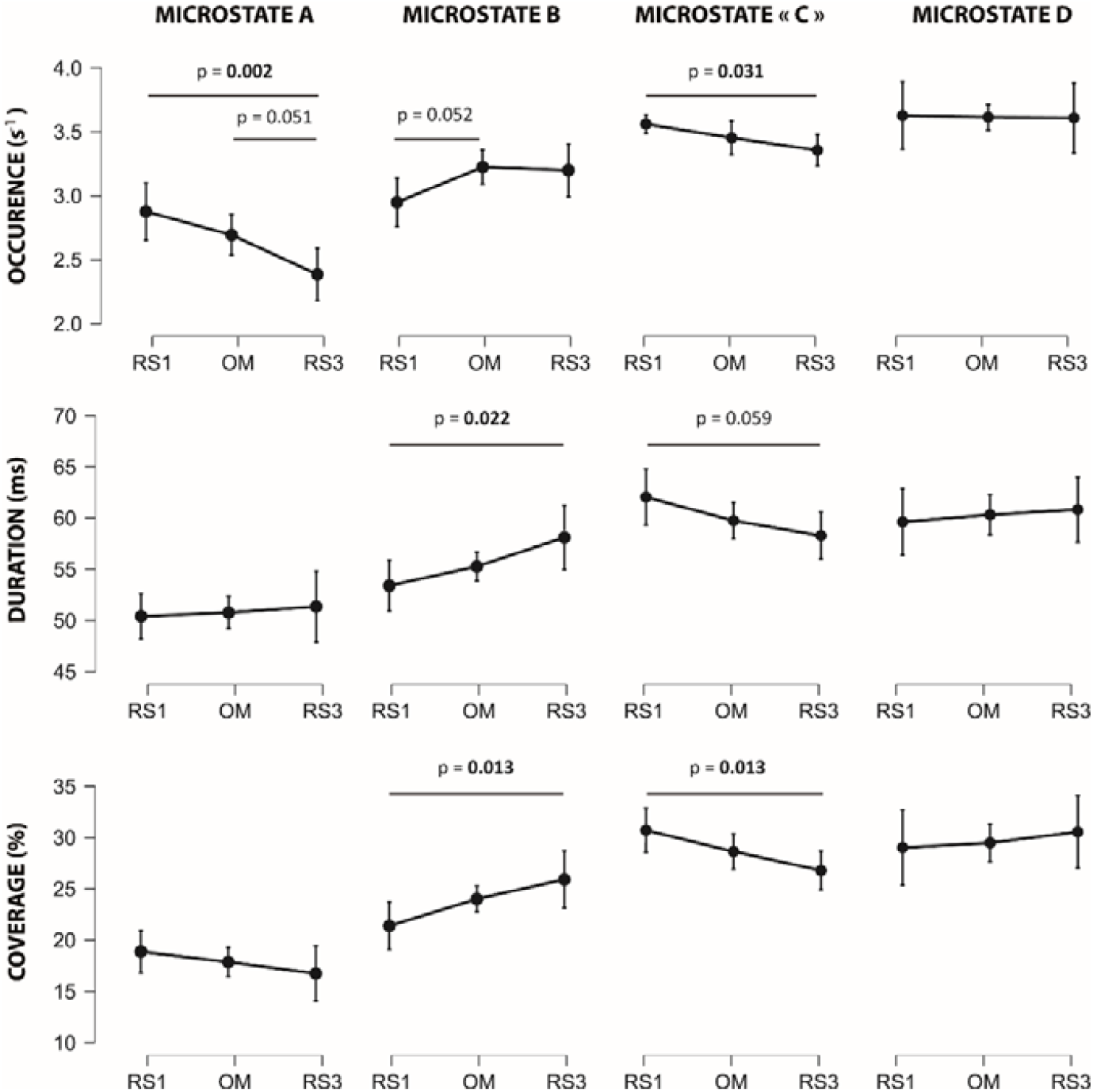
Microstates temporal parameters comparisons across periods within the meditator group. Error bars correspond to standard error.

### Microstate A

Repeated measure ANOVA revealed a period effect on the occurrence of microstate A (*F*(2,34) = 7.090, *p* = 0.003) in meditators. Post-hoc tests highlighted significant decrease between RS1 and RS3 (*t* = 3.725, *p* = 0.002) and between OM and RS3 (*t* = 2.339, *p* = 0.051) (Figure 3). No effect was found about the duration and the coverage of microstate A (All *F* < 1.130, *p* > 0.320).

Spearman’s rank correlation revealed that between OM and RS3, the decrease of the microstate A occurrence was negatively correlated to the increase in mind wandering (*ρ*(18) = -0.590, *p* = 0.009). The occurrence during RS3 (but not during RS1 and OM) were negatively correlated with the FFMQ total score (*ρ*(18) = -0.547, *p* = 0.023) in meditators, and not in non-meditators (*ρ*(18) = -0.236, *p* = 0.346) (Figure 4). This correlation in meditators was more specifically related to describing score of FFMQ (*ρ*(18) = -0.487, *p* = 0.048).

**Figure 4:**
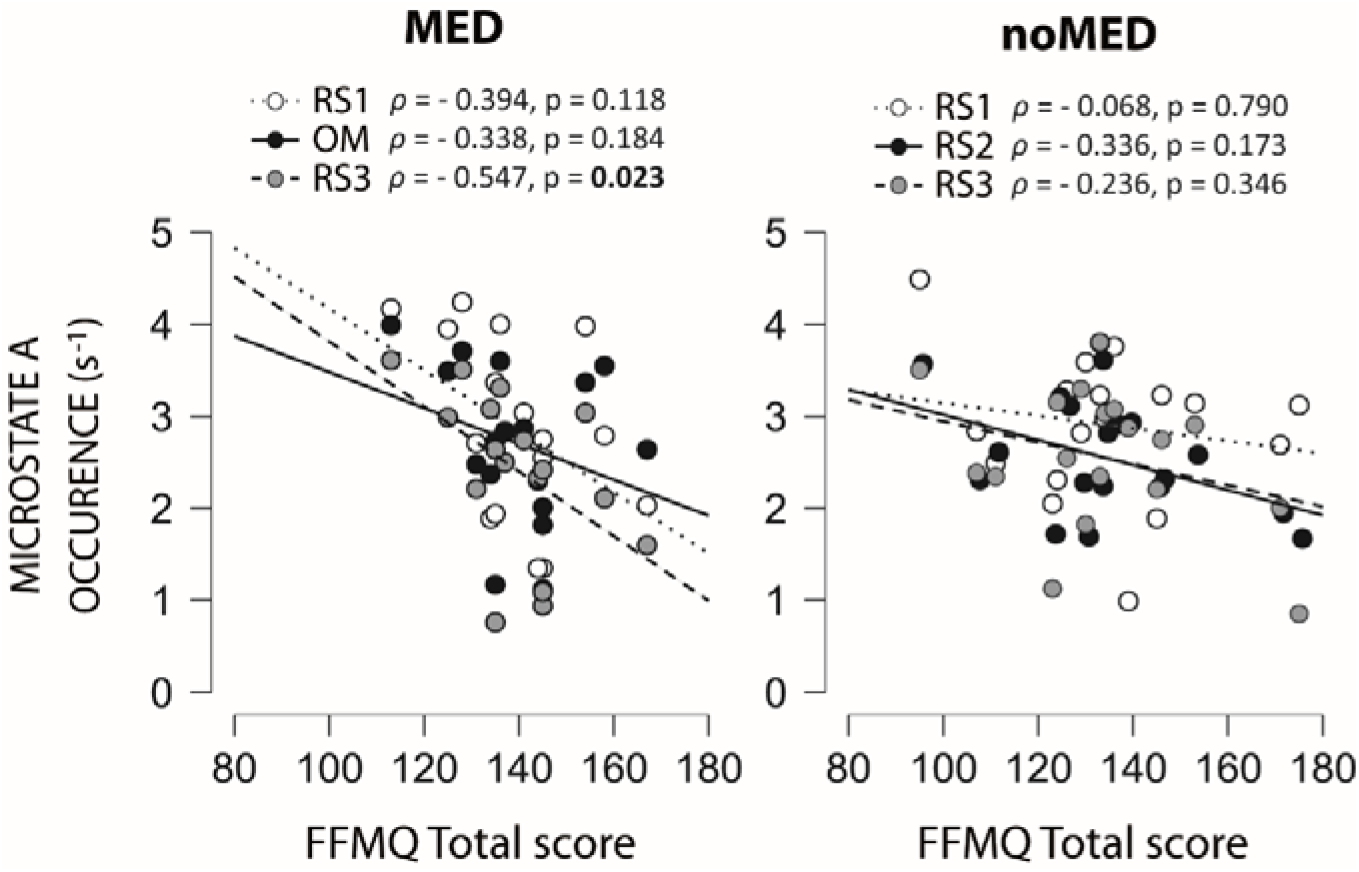
Correlations between the microstates A temporal parameters and the FFMQ total scores according to groups and periods.

### Microstate B

A period effect was found for microstate B duration (*F*(2,34) = 4.102, *p* = 0.025) and coverage (*F*(2,34) = 4.696, *p* = 0.016) in meditators. A trend for microstate B occurrence was also observed (*F*(2,34) = 3.224, *p* = 0.052). Post-hoc tests indicated significant increase in microstate B duration (*t* = -2.845, *p* = 0.022) and coverage (*t* = -3.052, *p* = 0.013) between RS1 and RS3 (Figure 3).

Spearman’s rank correlation revealed that the duration and coverage of microstate B in meditators were negatively correlated to the auditory distractibility during RS1 (*ρ*(18) = -0.474, *p* = 0.047; *ρ*(18) = -0.502, *p* = 0.034 respectively). Moreover, the increase of the duration and the coverage between RS1 and RS3 was positively correlated to the FFMQ non-reactivity scores (*ρ*(18) = 0.686, *p* = 0.002; *ρ*(18) = 0.736, *p* < 0.001 respectively). These increases appeared more specifically between OM and RS3 (*ρ*(18) = 0.565, *p* = 0.018; *ρ*(18) = 0.627, *p* = 0.007 respectively), and were negatively associated to increases in emotional charge (*ρ*(18) = -0.465, *p* = 0.052; *ρ*(18) = -0.526, *p* = 0.025 respectively). The duration and coverage of microstate B during RS3 were also positively correlated with FFMQ non-reactivity score in meditators (*ρ*(18) = 0.664, *p* = 0.004; *ρ*(18) = 0.658, *p* = 0.004 respectively), and not in non-meditators (*ρ*(18) = -0.006, *p* = 0.980; *ρ*(18) = -0.154, *p* = 0.540 respectively) (Figure 5).

**Figure 5:**
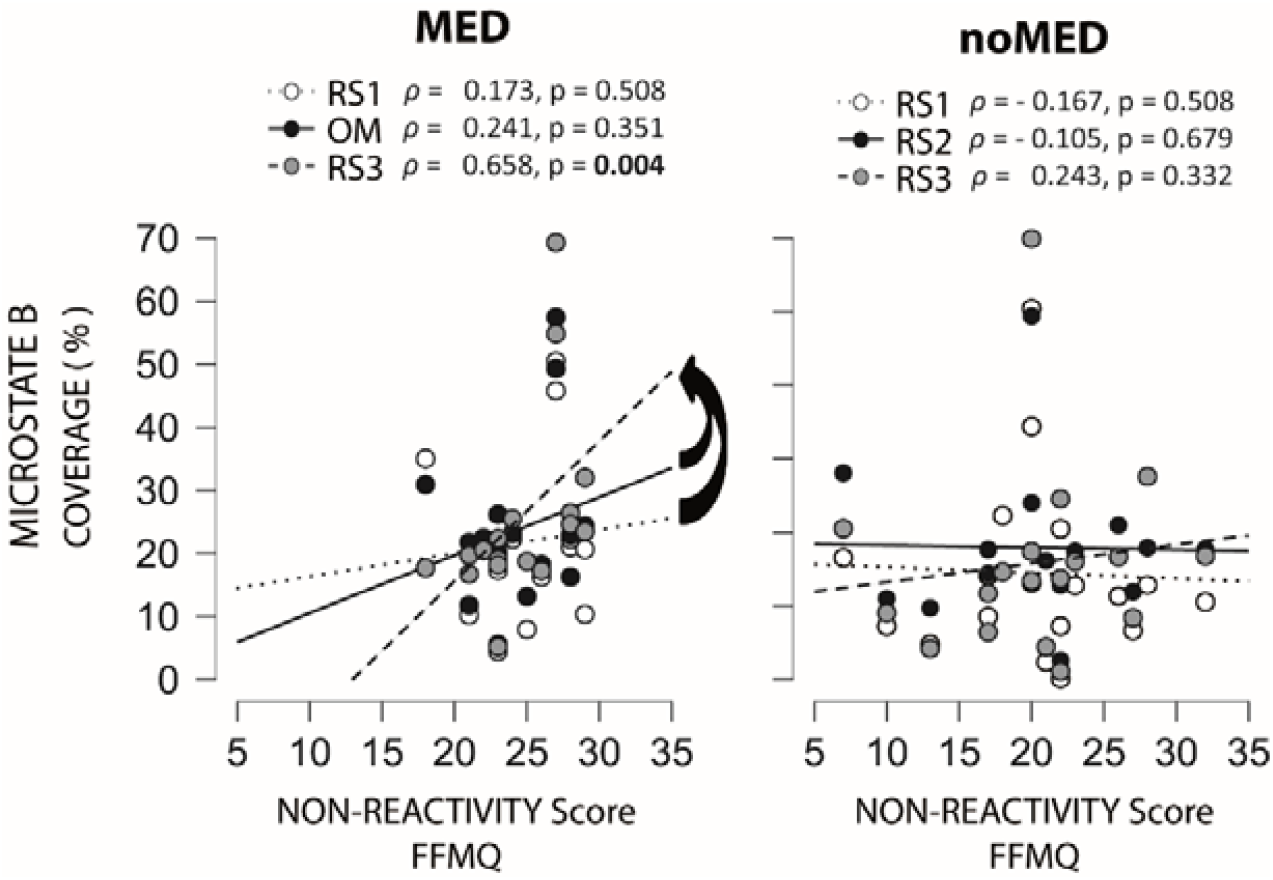
Correlations between the microstates B temporal parameters and the FFMQ non-reactivity scores according to groups and periods. The arrows indicate that increases between RS1 and RS3, as well as between OM and RS3 were positively correlated to non-reactivity scores (*ρ(18*) = 0.736, *p* < 0.001, *ρ*(18) = 0.627, *p* = 0.007 respectively).

The increase of the occurrence and coverage between RS1 and OM were correlated to the FFMQ total scores (*ρ*(18) = 0.581, *p* = 0.014) and FFMQ observing scores (*ρ*(18) = 0.509, *p* = 0.037) respectively. Positive correlation was also found between coverage of microstate B during OM and FFMQ observing scores (*ρ*(18) = 0.507, *p* = 0.038) in meditators, and not in non-meditators (*ρ*(18) = -0.333, *p* = 0.177).

### Microstate C

Microstate C showed significant differences in occurrence (*F*(2,34) = 3.691, *p* = 0.035) and coverage (*F*(2,34) = 4.638, *p* = 0.017) between periods in meditators. A trend was also observed for duration (*F*(2,34) = 3.043, *p* = 0.059). Post-hoc tests indicated significant decrease in microstate C occurrence (*t* = 2.715, *p* = 0.031) and coverage (*t* = 3.044, *p* = 0.013) between RS1 and RS3 (Figure 3).

Spearman’s rank correlation revealed that the occurrences of microstate C in the three periods (RS1, OM, RS3) were negatively correlated to FFMQ non-reactivity scores, being more significant during OM (RS1: *ρ(18)* = - 0.536, *p* = 0.026; OM: *ρ(18)* = - 0.660, *p* = 0.006; RS3: *ρ(18)* = - 0.528, *p* = 0.029) in meditators, but not in non-meditators (RS1: ρ(18) = - 0.299, *p* = 0.229; RS2: *ρ(18)* = - 0.121, *p* = 0.631; RS3: *ρ(18)* = - 0.112, *p* = 0.658) (Figure 6). Moreover, the decrease of the microstate C occurrence between RS1 and OM in meditators was positively correlated to the FFMQ non-reactivity scores (*ρ(18)* = 0.567, *p* = 0.018) and was negatively correlated to changes in mind wandering (*ρ(18)* = - 0.546, *p* = 0.019) and in emotional charge (*ρ(18)* = - 0.500, *p* = 0.034). Microstate C occurrence were negatively correlated to the emotional charge during OM in meditators (*ρ(18)* = - 0.522, *p* = 0.026), but also during RS2 in non-meditators (*ρ(18)* = - 0.487, *p* = 0.040). During RS3, microstate C occurrence and coverage were negatively correlated to mind wandering only for meditators (*ρ(18)* = - 0.606, *p* = 0.008; coverage: *ρ(18)* = - 0.531, *p* = 0.023 respectively) (Supplementary Files 6).

**Figure 6:**
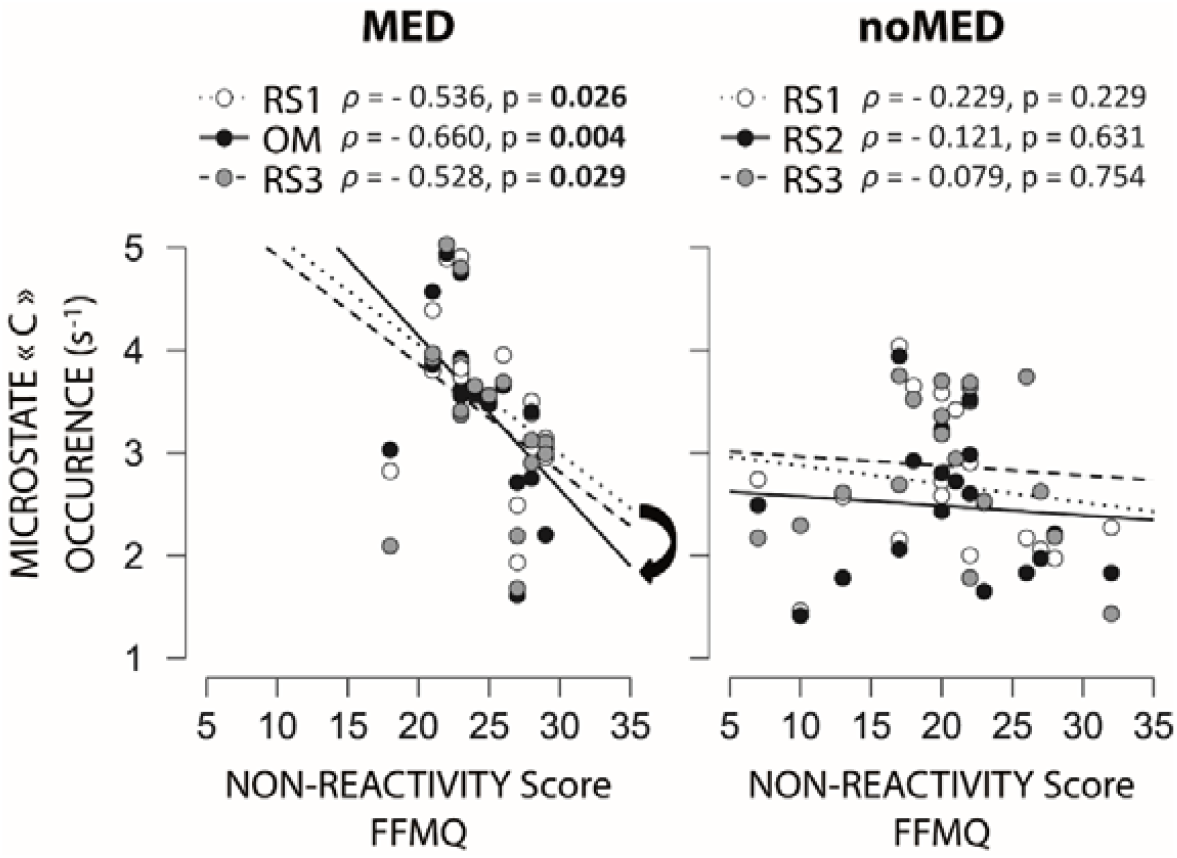
Correlations between the temporal parameters of microstate « C » and the FFMQ non-reactivity scores according to groups and periods. The arrow indicates that decrease between RS1 and OM were correlated to non-reactivity scores (*ρ(18)* = 0.567, *p* = 0.018).

In line with the trait analysis, we also found positive correlations between the duration and the coverage of microstate C and auditory distractibility during RS1 in non-meditators (*ρ(18)* = 0.602, *p* = 0.008; *ρ(18)* = 0.549, *p* = 0.018 respectively) and in a lesser extend in meditators (*ρ(18)* = 0.513, *p* = 0.029; *ρ(18)* = 0.417, *p* = 0.085 respectively).

### Microstate D

No changes were found for microstate D temporal parameters between periods for meditators (all *F* < 0.280, *p* > 0.678) (Figure 3). Coverages of microstate D during RS1 and OM were positively correlated to non-judgment facet of FFMQ in meditators (RS1: *ρ(18)* = 0.620, *p* = 0.008; OM: *ρ(18)* = 0.564, *p* = 0.018; but not for non-meditators: RS1: *ρ(18)* = 0.381, *p* = 0.118; RS2: *ρ(18)* = 0.147, *p* = 0.560). More specifically, these correlations were respectively sustained by duration during RS1 (*ρ(18)* = 0.518, *p* = 0.033) and occurrence during OM (*ρ(18)* = 0.559, *p* = 0.020).

### SOURCE LOCALIZATION ANALYSIS

Our results of the microstates characterization in the state stream analysis have revealed that microstate « C » differentiated meditators and non-meditators for the RS1 and RS2/OM periods, and the three periods for non-meditators groups. Figure 7 illustrates the statistical maps of the source analysis of microstate « C » topographies compared between groups (upper part). In RS1, higher activity in the right hemisphere including retrosplenial cortex (BA30: 28.3, -46.6, 9.4), PCC (BA31: 19.1, -65.1, 16.9), and a part of cuneus (BA18: 19.2, -67.1, 15.8) in meditators compared to non-meditators. In contrast, non-meditators showed higher activity in the left hemisphere area peaking in the superior temporal gyrus (BA22: -50.8, 14.4, -0.9) compared to meditators (Figure 7, upper part, on the left). In RS2, microstate « C » showed local involvement of the right cerebellum (culmen: -10.1, -63.3, 9.5; declive: 10.3, -71.5, 17.3) and bilaterally in cuneus (BA18 L: -10.1, -78.1, 23.1; R: 11, -86.6, 14.1) in meditators with respect to non-meditators. Sources of microstate « C » in non-meditators in RS2 showed distributed nodes of activities localized in the supramarginal gyrus (BA40: -39.1, -51.7, 34.6), ventral premotor cortex (BA6: -32.2, 3.4, 23.0) of the left hemisphere, as well as in dorsal premotor cortex (BA6: 36.9, -6.3, 48.5) and dlPFC (BA9: 36.4, 3.7, 31.6) in the right hemisphere. Concerning RS3, no significant generator activity was highlighted between groups except for a marginal contribution of the left hemisphere in a localized fontal single node (BA8: -21.7, 20.5, 32.6) in non-meditators (Figure 7).

**Figure 7:**
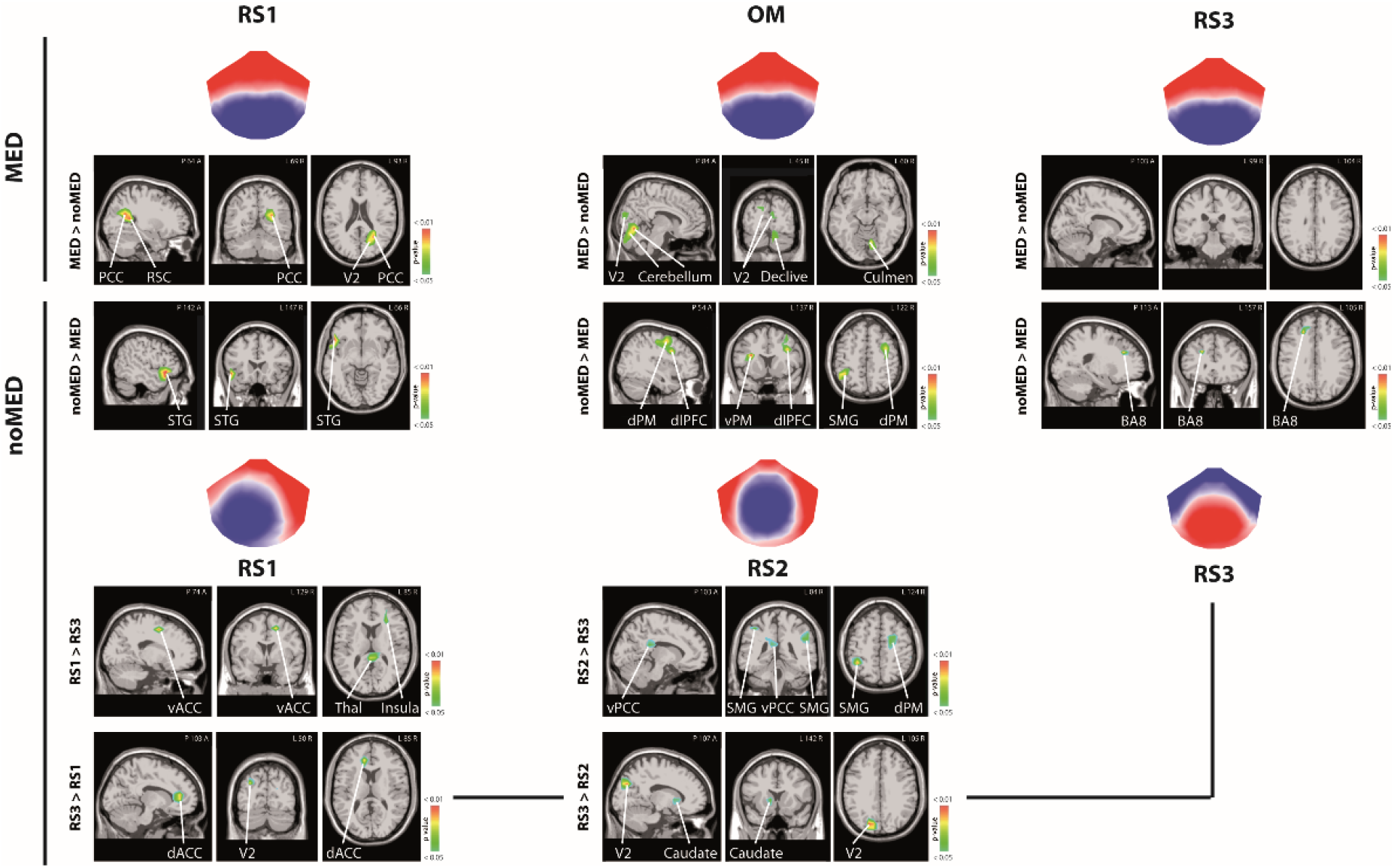
Statistical maps of microstate « C » between groups and periods. d/vACC: dorsal/ventral anterior cingulate cortex, PCC: posterior cingulate cortex; dlPFC: dorsolateral prefrontal cortex; d/vPM: dorsal/ventral premotor cortex; RSC: retrosplenial cortex; STG: supramarginal gyrus; V2: secondary visual cortex; BA8: Broadmann area 8, a part of frontal cortex including frontal eyes fields.

Additionally, we explored the statistical maps of the significant sources accounting for microstate « C » in RS1 and RS2 compared to RS3 within non-meditators (Figure 7, lower part), as we previously found high correlations of maps « C » between groups for this period (see above). Sources of microstate « C » were higher in the thalamus (pulvinar: 5.4, -36.2, 9.6), ventral ACC (BA24: 16.2, -6.0, 47.8), and insula (25.8, 26.0, 15.7) in the right hemisphere in RS1 than in RS3. Left dorsal ACC (BA32: -14.1, 31.8, 13.8) and bilateral cuneus (BA19 L: -28.2, -78.8, 24.1; BA18 R: 19.2, -68.1, 15.7) were higher in RS3 than RS1 period. Sources in RS2 presented higher activities than RS3 in supramarginal gyrus (BA40 L: -39.1, -51.7, 43.6), ventral premotor cortex (BA6: -42.2, 1.4, 29.2) and ventral PCC (BA23: -9.5, -36.7, 18.7) in left hemisphere, as well as supramarginal gyrus (BA40 R: 47.8, -36.7, 27.2), angular gyrus (BA39: 55.1, -53.5, 14.9) and dorsal premotor cortex (BA6: 27.1, -7.4, 49.3) in right hemisphere. RS3 showed higher activities in left cuneus (BA18: -18.1, -79.6, 24.8) and left caudate (body: -12.3, 6.0, 5.1) than RS2 (Figure 6).

## DISCUSSION

In the present study, we investigated the microstates dynamics induced by OM, dissociating mindfulness state effects from traits effects. Questionnaire scores have shown that meditators presented higher traits related to observing and non-reactivity to inner experiences than non-meditators, both features underlying OM practice. Trait microstate analysis revealed a composition of 4 canonical microstates (labeled A to D) explaining 70.18 % of the recorded data at rest across all participants. We found a negative correlation between the FFMQ total score and temporal parameters of microstate A (occurrence, coverage). Considering FFMQ sub scores, these parameters were negatively associated with the non-reactivity and non-judgment facets of mindfulness. The FFMQ observing and non-reactivity scores were also negatively correlated to the temporal parameters of microstate C (occurrence, duration, and coverage). According to mindfulness facets defined by FFMQ [Baer et al., 2008], these data suggest that while microstate A and C seem to be linked to reactive behaviors, microstate A occurs about an evaluative mental state toward thoughts and feelings, while microstate C is related to an inattentive mental state toward (internal or external) sensory information underlying present moment experience.

Regarding state analysis, the same four trait-related microstate topographies remained stable across periods, explaining about 72.1% of the data, for meditators. Compared to the resting state before meditation, specific changes were found in the temporal parameters of microstate A (occurrence), B (duration, and coverage), and C (occurrence and coverage) during and after OM in meditators. Namely, the occurrence of microstate A decreased after OM. The duration and coverage of microstate B increased after OM in particular in meditators showing high FFMQ non-reactivity scores. Finally, the occurrence and coverage of microstate C decreased after OM and were negatively correlated to non-reactivity scores regardless of periods (and more especially during OM). These results suggest that beyond trait-related effects, the practice of OM decreases the occurrence of microstates A and C reflecting decreased mental state reactivity, while the increases of the microstate B temporal parameters would rely on attention stability towards the present moment experience.

In this line, microstate A has been associated with the primary auditory cortex (BA41), Wernicke area (BA22), and insula activities [Custo et al., 2017] and was generally attributed to audio-phonological processing [Britz et al., 2010]. In this context, the occurrence of microstate A could index processes related to ruminations, which mindfulness aims to mitigate. Interestingly, a recent report indicated that the occurrence of microstate A is positively correlated with depression severity, where ruminations are highly present [Damborská et al., 2019]. Also, a previous study on expert meditators reported a similar decrease in the occurrence of microstates A and C during transcendental meditation [Faber et al., 2017], consisting of mentally repeating a series of unmeaning words [Travis and Parim, 2017]. Their results showed that the meditation transcending phase was marked by lower microstate A and C occurrences compared to undirected mentation (i.e. mind wandering), characterizing a decreased salience of internally generated thought (i.e. more detached and less evaluative processing) [Faber et al., 2017]. Here, we showed that microstate A occurrence decreased, in particular in meditators reporting a low increase in mind wandering after OM. Moreover, microstate A occurrence after OM was negatively correlated to FFMQ total score in meditators, and more specifically with the describing facet referring to the ability to label internal experiences with words [Baer et al., 2008]. According to Faber et al. [2017], these results suggest that the influence of OM on microstate A relies on the metacognitive ability to identify experiences beyond ruminations.

Microstate B has been related to the visual system [Britz et al., 2010; Seitzman et al., 2017] and reflects the activity of the cuneus (BA17, 18) and secondarily in the insula, right claustrum, and frontal eyes fields [Britz et al., 2010; Custo et al., 2017]. A previous case study showed a longer duration of microstate B during meditation in expert meditators [Faber et al., 2005]. Here, we found an increased duration and coverage of microstate B after OM in meditators, in particular in those with high non-reactivity scores. Interestingly, we also observed an association between the increase of microstate B coverage during OM and the observing facet of mindfulness. Our subjects being eyes closed during each period, the increase of microstate B temporal parameters might reflect working memory processes which are considered the core cognitive mechanism of vivid perceived experience during mindfulness meditation [Jha et al., 2019; Lutz et al., 2015; MacLean et al., 2010]. On the other hand, our data did not show a significant influence of OM on microstate D temporal parameters which has been associated with attention control [Britz et al., 2010; Milz et al., 2016] and with context updating of mental content [Brodbeck et al.0, 2012; Katayama et al., 2007; Kindler et al., 2011; Rieger et al., 2016] both underlying mindfulness meditation. One possible explanation could be provided by previous studies suggesting that attention control and context updating have opposite effects on microstate D parameters. Indeed, microstate D parameters have been reported lowered during goal-directed attention [Milz et al., 2016], while they would be raised by context updating [Rieger et al., 2016].

The main result of our study concerned microstate C which was closely related to the non-reactivity facet of mindfulness and whose occurrence decreased after OM. It has been proposed that microstate C was linked to interoceptive-autonomic processing involved in the integration of salient emotional information [Britz et al., 2010]. Microstate C has been associated with two major interacting hubs of DNM, namely the ACC and the PCC [Michel and Koenig, 2018]. The dynamic of microstate C would reflect the activity of the SN playing a critical role in switching between central-executive functions and the default mode [Britz et al., 2010]. The SN largely overlaps with the ventral attentional system involved in detecting and orienting the organism toward salient external or internal events in a bottom-up fashion [Corbetta and Shulman, 2002]. Here, we found associations between the duration of microstate C, the auditory distractibility ratings, and the FFMQ observing scores across all participants during the first recording at rest. While the two first were positively associated, they were both negatively correlated to the FFMQ observing score. These data seem to confirm that microstate C reflects SN activity, and suggest that SN activity was modified at rest in participants with a higher tendency to pay attention to the sensory information of lived experience. We found further that temporal parameters of microstate C were negatively correlated to the non-reactivity facet of mindfulness. Accordingly, meditators showed lower temporal parameters of microstate C at rest than non-meditators, suggesting that the SN underlies the tendency to allow thoughts and feelings to come and go, without getting caught up in or carried away by them.

Interestingly, the cluster decompositions provided by state analysis revealed dissimilar maps « C » between groups at RS1 and RS2/OM, while the other canonical maps A, B, and D remained similar between groups. Although topographically dissimilar, temporal parameters of both microstates « C » were correlated to distractibility and emotional charge ratings of their respective groups, confirming that these microstates « C » shared common salience processes. In this context, meditators reported lower emotional charges than non-meditators at RS1 and RS2/OM. This could indicate the activation of emotion successful integration networks [Britz et al., 2010] specifically in meditators explaining the clustering identification of microstate C. In fact, topographies for meditators were stable across recording periods whereas non-meditators present differences between the three periods. Studies suggest a high sensibility of microstates to emotions [Chen et al., 2021] which are viewed as products at the brain network level [Lindquist et al., 2012]. In particular, the DNM plays a key role in the construction of discrete emotional experiences [Satpute and Lindquist, 2019]. In this emotional context, the lower GEV of non-meditators’ microstate compositions (Figure 1), could reflect higher variability in the temporal dynamics of brain activity in this group compared to meditators. This higher variability could have induced different topographic identifications provided by fixed *k*-means clustering. In contrast, previous studies suggested that attentional control underlying meditation induced lower variability of brain network dynamics through a decreased PCC connectivity in meditators [Panda et al., 2016]. Considering also reports showing that mindfulness uncouples brain areas related to sensory, affective, or evaluative aspects of lived experience [Grant et al., 2011; Zorn et al., 2020], it is, therefore, possible that mindfulness leads specific brain sub-networks to work more stably.

At rest during the first recording, source analysis showed that the microstate C topography was characterized by the higher contribution of PCC in meditators while it was characterized by contributions of right superior temporal gyrus (BA22, related to non-verbal sound processing), as well as the ventral anterior cingulate cortex (vACC), insula, and thalamus (three major nodes of the SN [Seeley, 2019]) in non-meditators. In this line, Custo et al. [2017] highlighted that microstate C could collapse into two microstates (C and F in their study): one related to the more posterior part of the DMN, and the other being related to the SN. It has been suggested further that these two microstates reflect anti-correlated network activities [Zanesco et al., 2021a]. Here, the topographic dissimilarity of microstate C between groups at rest seems to confirm the less prominent activity of the SN in trained meditators. A recent meta-analysis of fMRI studies indicated that grey matter changes in the insula (and ACC) are the most consistent effect observed across mindfulness studies [Pernet et al., 2021]. A previous study by Bréchet et al. [2021] also reported topographic changes in the clustering identification of microstate C at rest after 6-week meditation training in novice meditators. Using source localization analysis, they revealed that the microstate C after training was carried by higher activity bilaterally in the insula and the left supramarginal gyrus (BA40) which they interpreted as a reorganization of functional connectivity in fronto-insular-parietal networks after training.

During the second recording, microstate C at rest in non-meditators was characterized by widespread activities including dlPFC, supramarginal gyrus, and ventral PCC which are described in internally directed thoughts and self-generated emotions [Damasio et al., 2000; Kropf et al., 2019; Leech and Sharp, 2014]. In contrast, microstate C during OM in meditators was characterized by a strong activity of the cerebellum (anterior and posterior part). In parallel, the occurrence of microstate C decreased, in particular in meditators showing high FFMQ non-reactivity scores. According to the critical role of the cerebellum in behavioral control and learning [Cheron et al., 2013; Cheron et al., 2016; De Zeeuw, 2021], this structure might represent a major neural substrate of non-reactivity behavior (motor, affective and cognitive inhibition), underlying mindfulness meditation. In a previous study, we have recently suggested that the anterior part of the cerebellum contributes to automatic features of sensorimotor inhibition control by integrating into its multiple internal representations the appropriate behavioral responses to stimuli [Zarka et al., 2021]. The role of its posterior part in affective and (non-motor) cognitive functions is also largely recognized [Argyropoulos et al., 2020; Stoodley and Schmahmann, 2018]. According to Schmahmann (2019), the cerebellum is an oscillation dampener smoothing out motor but also emotional and cognitive performance. It modulates behaviors by comparing the consequences of (non-)actions with the intended outcome, matching reality with perceived reality. In other words, the cerebellum could represent the essential neural substrate of cognitive defusion mechanisms underlying mindfulness meditation [Zorn et al., 2021] and associated salutary changes in the brain’s working memory system [Jha et al., 2019; Stoodley and Schmahmann, 2009; Stoodley and Schmahmann, 2018].

## CONCLUSION

To conclude, this study aimed to characterize microstate dynamics induced by non-reactive attention meditation using a multimodal approach. Our results revealed that the mindfulness traits were negatively correlated with temporal parameters of microstates A and C at rest. We showed further that OM decreases the occurrence of microstates A and C reflecting decreased mental state reactivity, while it increases the duration of microstate B through attention stability towards the present moment experience. While the decreased occurrence of microstate A would be related to ruminations weakening, decreased microstate C occurrence was associated with a modified activity of SN and might represent an index of interoceptive processing. These findings strongly encourage more research to assess the use of the microstate temporal parameters as a biomarker of the SN (re-)activity, as well as objectify the brain changes induced by non-reactive attention training and their salutary effects on mental health.

## Data Availability

All data produced in the present study are available upon reasonable request to the authors

## DATA AVAILABILITY STATEMENT

All relevant data will be available from the corresponding authors upon reasonable request.

## CONFLICT STATEMENT

The authors declare that the research was conducted in the absence of any commercial or financial relationships that could be construed as a potential conflict of interest.

## ACKNOWLEDGMENTS

This work was funded by the Université Libre de Bruxelles (Belgium), the Secretaria Nacional de Ciencia y Tecnologia (Senescyt, Ecuador) and the Fonds G. Leibu. We warmly thank L. Felz, F. Bauwens, C. Maskens who kindly took part in this research as mindfulness expert, as well as I. Kotsou for expert scientific advice. We also wish to express our gratitude to the participants of the study, as well as our thanks to E. Pecoraro, E. Hortmans, E. Toussaint, and T. D’Angelo for essential administrative and technical assistance.

## Notes

### Competing Interest Statement

The authors have declared no competing interest.

### Funding Statement

This work was funded by the Universite Libre de Bruxelles (Belgium), the Secretaria Nacional de Ciencia y Tecnologia (Senescyt, Ecuador) and the Fonds G. Leibu.

### Author Declarations

Ethics committee of Erasme Hospital (ULB, 021/406) gave ethical approval for this work.

